# Pushing the boundary of child well-being: A spatial examination of child mortality in transition zones of extreme economic inequality and material hardship

**DOI:** 10.1101/2024.12.02.24318335

**Authors:** Gia Elise Barboza-Salerno, Brittany Liebhard, Karla Schockley-McCarthy, Sharefa Duhaney

## Abstract

How do patterns of socioeconomic inequality shape the risk of child fatality in urban areas? Studies have demonstrated that intentional and accidental deaths of children are highly clustered into areas of social disadvantage. However, in complex urban settings, the risk of death to children is likely to exhibit a more localized spatial structure characterized by rapid changes in child fatality risk. The present research uses Bayesian hierarchical modeling to detect spatial discontinuities in child fatality risk in transition areas defined by elevated levels of economic hardship and inequality (EHI). The analysis detected 413 neighborhood boundaries characterized by extreme differences in EHI (i.e., a difference of four deciles). Living in proximity to a boundary of extreme difference, called a social frontier, is associated with a 22% higher relative risk of child fatality beyond measures of neighborhood racial segregation, concentrated disadvantage, residential mobility, and immigrant concentration. The significance of identifying neighborhoods characterized as a social frontier where children may benefit from additional preventive interventions is discussed in context.

## Introduction

Child mortality is a critical public health issue in the United States. In 2021, the all-cause child mortality rate reached its highest level since 2008, with significant increases observed during the COVID-19 pandemic [1–2]. In Cook County, Chicago, the geographic location of the present study, 599 children under the age of 18 died in 2022, a 15.9% increase from the previous year [3–4]. These figures mask substantial heterogeneity in child mortality across race, socioeconomic status, and geography. More than half (56.6%) of all deaths to children under 18 years old occurred in the city of Chicago. Cook County exemplifies the national trend of Black children disproportionality in mortality rates, with non-Hispanic Black children making up 61.5% (1369) of all child fatalities between 2015 and 2023 while comprising only 23.6% of the child population. On the other hand, non-Hispanic White children were significantly underrepresented compared to their share of the population, comprising 16.2% of the deaths but 65.1% of children [5].

Significant disparities in child mortality rates across regions and socioeconomic groups indicate that economic inequality amplifies existing racial and ethnic disparities. Geographic location is a key factor in child mortality, influencing access to healthcare, the quality of public health infrastructure, and the extent of socioeconomic challenges. Urban areas with high levels of poverty are particularly vulnerable, with children facing increased risks of mortality due to inadequate housing, environmental hazards, and limited access to quality healthcare. Low-income families are disproportionately affected, as children in these households often experience poor health outcomes driven by factors such as insufficient nutrition [6–8], limited access to healthcare [9–11], and greater exposure to environmental toxins [12–13]. These conditions contribute to a higher prevalence of adverse health outcomes, including preterm births, low birth weight, and chronic illnesses, all of which significantly elevate the risk of infant and child mortality [14].

The existing literature on child mortality disparities across income levels largely focuses on individuals or households, often neglecting the role of neighborhood-level economic inequality [15–16]. While it is well-documented that child mortality rates vary geographically and are influenced by socioeconomic factors, no study to date has directly examined how differences in economic inequality between neighborhoods relate to child mortality [17]. This study addresses that gap by investigating how area-level income gradients—particularly those marked by sharp socioeconomic differences between adjacent neighborhoods—affect child mortality rates. Specifically, we aim to determine whether spatial discontinuities, defined as sudden shifts in economic inequality between neighborhoods, contribute to an increased relative risk of child mortality. Our analysis quantifies the impact of these spatial inequalities on child mortality risk, extending beyond traditional metrics of social vulnerability.

### Economic inequality and child mortality

During the past four decades, income inequality in the Chicago region increased by nearly 36%, surpassing the 19% rate of increase for the United States [18]. Socioeconomic inequality in mortality and morbidity, including a lack of secure housing, job opportunities, quality education, healthcare, and childcare, remains a critical social determinant of mortality among youth [16,19]. Spatial concentrations of child mortality risk have been attributed to variations in material wealth across communities [19]. Despite significant declines in child mortality rates since 1950, socio-economic disparities in child mortality across levels of socio-economic disadvantage have not only persisted [20], they have increased [21]. Past research has demonstrated socioeconomic gradients in all-cause child mortality and specific-cause mortality in vehicular accidents [22], pedestrian injury [23], fires [24–25], drowning [26], sudden infant death syndrome [27], child maltreatment [28], and congenital conditions [29]. The survival advantage of children who live in the least deprived neighborhoods has been attributed to the differences in quality education, employment, and healthcare opportunities found in wealthier areas [20,30]. Not all socio-economically disadvantaged neighborhoods within the same geographic region experience the same level of inequality, however. In metropolitan areas, the observed spatial heterogeneity in child mortality outcomes is influenced by a diverse range of social, economic, and environmental determinants that vary widely depending on the region being considered [19,31,32, 33]. Neighborhood structural disadvantage has been reinforced by punitive justice-related, discriminatory housing policies and antipoverty policies that aim to supervise and adjudicate family life in impoverished areas that serve as pathways by which structural determinants shape health outcomes for children [34–35].

### Neighborhood boundaries as social frontiers

Neighborhood boundaries may be defined as homogenous areas, such as census tracts or counties, or as spatial zones of rapid change called social frontiers [36–38]. Neighborhoods that exhibit internal similarity based on the specified characteristics may differ due to their sociospatial position. Social frontiers, which represent spatial divisions in racial, ethnic, religious, cultural, or social characteristics that separate neighboring communities, act as “cliff edges” between adjacent neighborhoods [36]. Boundaries are locations where socio-economic status changes rapidly and are of scientific interest because they represent zones of dynamic geographic change that distinguish social factors on one side of the boundary from those on the other [39]. Also, social frontier boundaries are open, meaning they appear as line segments (i.e., not polygons) and do not enclose an area. Because the processes that give rise to boundaries are not always associated with homogeneous areas, social frontiers may matter more on “one side where they divide Black [*sic*] city residents from surrounding White [*sic*] suburban areas, but they may not matter on another side (p. 20)”[40].

Social frontiers typically coincide with the edges of highly segregated regions where neighboring areas are more homogenous [40–43]. Social frontiers may be caused by a “strong aversion [among residents] to living at the interface of communities in conflict (pg. 274)” [36,44] or because of “benign forces” [36,45] and unintended consequences of micro-level (i.e., individual) decisions. Alternatively, social frontiers may emerge from explicit and intentional policies that have prevented people in lower-income communities of color from accumulating wealth and investing in home ownership rather than be an “accident of economic circumstance, demographic trends, personal preference, and private discrimination” [46]. Regardless of their origins, social frontiers are places of stark spatial divisions with complex social consequences. Such variation has important implications for understanding the impacts of socio-economic segregation and its disparate effects on health and well-being. These boundaries demarcate areas with differences in level of service and resource distribution, cultural and place-based identity, and social interactions, all of which can impact the social dynamics of these areas [47]. Residents’ social interactions and demographic characteristics shape the meaning and perception of the places where people live, even within the same geographic area [48].

### How do social frontiers affect child mortality risk?

To fully understand the impact of spatial opportunity structure on health and well-being, it is essential to look beyond the characteristics and interactions within individual neighborhoods [47,49]. More specifically, it is important to also consider spillover effects—how conditions in one neighborhood influence the health and well-being of surrounding neighborhoods [47]. Externality space theory provides an organizing framework for conceptualizing differences across bordering areas. According to the theory, places “where individuals do not share externalities with proximate others is axiomatically a boundary between … neighborhoods … that identify schisms in the social meaning or attributes of space at a given point … (pg. 83).” Boundary areas significantly alter the benefits and detriments that individuals derive from a particular location [47].

An externality space consists of three key dimensions: congruence, generality, and accordance. *Congruence* refers to the level of concordance between the externality space and a defined geographic boundary. In the present case, the congruence between EHI and child mortality risk is high when confined to specific neighborhoods. The *generality* of an externality space refers to how consistently an individual’s well-being is mapped to the same area. As applied here, if externalities about neighborhood economic inequality (e.g., healthcare access) map to increases in child mortality risk, then the externality space has high generality. *Accordance* refers to whether all individuals share a common understanding about where boundaries of EHI begin and end (i.e., the externality space). The externality space theory of neighborhood boundaries introduces a novel approach to understanding urban dynamics [47] because it emphasizes important linkages across different sociospatial contexts— including the interrelatedness between space, sociability, and spatial conditions [35]. In short, this perspective calls attention to residents’ social marginalization due to the imposed nature of neighborhood physical boundaries.

At social frontiers, spatial proximity may decrease social interaction and the sharing of social institutions across groups [50]. Social frontiers may erect barriers to increased social interactions between neighbors, leading to greater mistrust and misunderstanding between groups, lower levels of collective efficacy, and increased child mortality [36,51]. This is supported by Legewie (2018), who found that even after considering traditional indicators of social disorganization, including residential instability, immigrant concentration, ethnic heterogeneity, and concentrated disadvantage, racial neighborhood boundaries continued to predict violent crime and homicide risk in Chicago. According to Legewie, boundary areas create opportunities for criminal behavior because of the lack of social control and cohesion characterized by adjacent homogeneous areas and the potential for intergroup conflict. Legewie (2018) used a technique known as ‘areal wombling’ to detect boundaries based on sharp changes in racial-ethnic composition. Drawing on multiple theories, he claims that “…boundaries lack the social control and cohesion of adjacent homogeneous areas; are contested between groups, provoking intergroup conflict; and create opportunities for criminal behavior” (Legewie, 2018, p 1958). In this study, violent crime was higher at boundary locations representing sharp delineations across racial-ethnic composition net of neighborhood characteristics, measures of spatial interdependence, and other physical boundaries. Kim and Hipp (2018) similarly found that nearness to the city boundary as a distance decay predicted increased motor vehicle theft [52]. Boundaries between two areas have been shown to increase crime opportunities, partly because of the possibility of fewer guardians, or the transitory nature of these locations, making escape quicker and easier for perpetrators of violent crime [53].

Regarding child mortality specifically, the perception of relative resource deprivation can lead to social disenfranchisement, mental distress, and poor health outcomes, particularly if the deprivation is attributed to social stigma and racism [50]. Research shows that the overlapping and reinforcing effects of residential segregation are associated with higher rates of infant mortality and preterm birth among Black infants [54,55]. One study found that the excess risk of mortality among preterm infants born in neighborhoods characterized by high relative concentrations of Black and poor residents is explained in part by the hospital in which they receive care [54]. More specifically, infants from predominately Black neighborhoods experienced worse outcomes if they were born in hospitals located in similar neighborhoods than if they were born in hospitals located in more White neighborhoods.

In contradistinction, studies have shown that spatial proximity can increase, rather than decrease, social interaction and the sharing of institutional resources across regions [50]. For example, ethnic diversity and multiculturalism have been viewed as positive externalities because they promote adherence to positive health behaviors consistent with child well-being through disease prevention and information about healthcare service locations [56–59]. Cross-cultural work has demonstrated locational advantages associated with proximity to affluent neighbors, such as greater accessibility to labor market opportunities and economic integration, but only when public spaces are shared effectively by socially dissimilar groups [34]. Moreover, increased social interaction in heterogeneous spaces can mitigate the impact of negative stereotypes and group threats and increase trust across groups [60,61]. In the context of child mortality, behavioral changes resulting from social interaction across diverse environments may be crucial for improving children’s survival. For instance, one study found that infant mortality is lower among rural-urban migrants than rural non-migrants, highlighting the role of social interaction in exposing individuals to new ideas, which can lead to changes in attitudes, lifestyles, and motivations [62].

### Current study

Past research confirms the importance of neighborhood boundaries as defining aspects of highly segregated urban areas, but the precise nature of the association remains challenging to predict. Living in proximity to a social frontier may impact child mortality by increasing access to buffering resources that can mitigate poor child health outcomes for those living in highly disadvantaged areas [63]. On the other hand, living near a social frontier may lead to social isolation and lower levels of collective efficacy, resulting in an increased mortality risk. To fully examine the effect of spatial inequality on child mortality, it is critical to consider multiple measures of economic inequality across and within regions. Boundary analysis captures the transition from low to high-class neighborhoods by detecting statistically significant latent discontinuities over a geographic surface [51,64]. In the present study, a novel form of spatial analysis called boundary detection is used to detect social frontiers of EHI - neighborhood boundaries based on extreme differences in economic inequality - and their association with child fatality in Cook County, Chicago. We hypothesized that living near a social frontier would be associated with decreased mortality due to the potential flow of resources from areas of less disadvantage.

## Methods

### Data

The Cook County Medical Examiner’s Office (CCMEO) provided data for this study. The CCMEA investigates all deaths that fall under various categories, including criminal violence, accidents, suicides, and deaths under suspicious or unusual circumstances. The CCMEO data includes detailed records of deaths reported in Cook County, Illinois, from August 2014 to the present. This data is updated daily and includes information such as the cause and manner of death, demographic details, and other relevant case information. We extracted all-cause mortality data for children under 18 from 2015-2023 for the study.

### Variables

#### Dependent Variable

##### Child Fatalities

Observed counts associated with each child death location, Y= (Y_1_,… Y_K_), were aggregated within census tracts. The number of children under 18 in each census tract in the county was downloaded from the American Community Survey (ACS) based on 5-year estimates for 2011 – 2015 and 2015 - 2019. Based on population size, the expected child fatality count, e= (e_1_,… e_K_), for each census tract was computed using the *SpatialEpi* [65] package in R.

#### Independent Variable

##### Economic Hardship and Inequality (EHI) Index

The EHI Index is a composite measure designed to assess the socioeconomic conditions of neighborhoods. This index is used to identify areas with high levels of economic deprivation and to understand the spatial distribution of economic hardship and inequality. It integrates multiple indicators to provide a comprehensive picture of economic hardship and inequality. The index includes income level, employment rate, housing stability, resource (healthcare, education, and transportation) access, and economic (income and wealth) distribution. We used the *get_adi* function in the *sociome* package in R to download the EHI index for 2015-2019. For more information about scale construction see [66].

#### Control Variables

Our models used multiple measures of social disorganization as control variables by creating indices corresponding to concentrated disadvantage, residential instability, immigrant concentration, and residential segregation similar to past research [51,67–69]. Following Legewie [51]: (1) concentrated disadvantage is a five-item index (eigenvalue 2.962; 59.2% variance) comprised of the following measures: poverty rate (factor loading: 0.847), unemployment rate (0.832), percentage of professional and management jobs (-0.681), the share of high-school graduates (0.616), and share single mother families (0.842); (2) residential instability is a three-item index (eigenvalue 1.85), 61.5% variance comprised of the percentage of renter-occupied units (0.998), share of residents who moved to another dwelling since 2005 (0.490), and housing unit rental vacancy rate (0.781); (3) immigrant concentration is a three-item index (eigenvalue 2.54, 84.7% variance) comprised of the share of foreign-born residents (0.913), the share of residents who speak English less than “very well” (0.998), and the share of Spanish-speaking residents (0.844). Further, to measure the level of racial or ethnic segregation, or concentration of different racial or ethnic groups, within each neighborhood, we computed the Hirschman-Herfindahl Index (HHI). The HHI is calculated by squaring the proportion of the population that belongs to each racial or ethnic group and then summing the squared values. This index ranges from 0 to 1, where a value closer to 0 indicates complete diversity (i.e., a more even distribution of racial or ethnic groups), and a value closer to 1 indicates a highly segregated neighborhood. All social disorganization variables were derived from the American Community Survey (ACS) 5-year estimates for 2015 – 2019 [70].

### Statistical Analysis

#### Overview of boundary detection approach

Our analysis proceeds as follows. We calculated the differences in EHI across adjacent areas using a spatial weights matrix. We examined the distribution of EHI differences between neighboring areas to identify geographic borders where these differences are statistically significant. Figure 1 illustrates a region characterized by a significant difference in EHI across boundaries.

**Figure 1.**
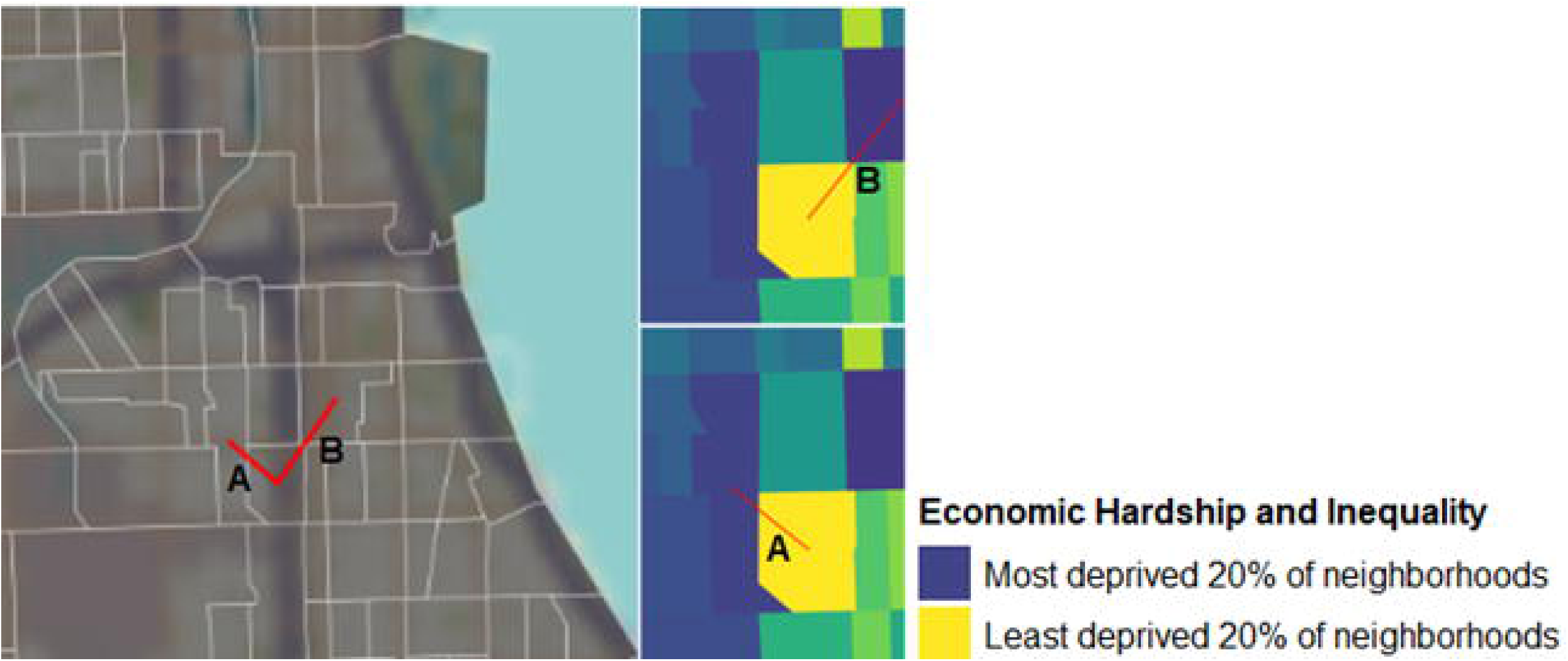
Illustration of the boundary detection method. The figure shows a census tract that neighbors two other census tracts labeled by lines A and B (left). The figure to the right shows that this tract was categorized as one of the least deprived 20% of neighborhoods by the EHI index, as shown in yellow. However, it borders two census tracts categorized as the most deprived 20% of census tracts by the EHI. Because this difference is greater or equal to four deciles, our analysis labels it a social frontier.

Next, we modeled the impact of these abrupt changes using locally adaptive spatial conditional autoregressive (CAR) models, as proposed by Lee [71] and Lee & Mitchel [72]. The dissimilarity metric identifies boundaries by computing the absolute difference in EHI deciles between pairs of census tracts. This model incorporates the spatial neighborhood matrix W to capture the absolute difference in EHI levels. To capture sharp distinctions in EHI across neighborhoods, we specified an absolute difference of at least four deciles, EHJ_kj_ = IEHJ_k_-EHJ_j_ I 2: 4. Setting the threshold at four deciles ensures we captured boundaries with pronounced economic differences while maintaining enough variation to detect meaningful relationships. Thereafter, we compared child mortality rates among pairs of contiguous census tracts joined by a boundary “frontier” against those not bordering a frontier to account for potential variations in the impact of neighborhood boundary differences.

#### Modeling

Following Lee and Mitchell (2012), a Bayesian spatial CAR model was specified as

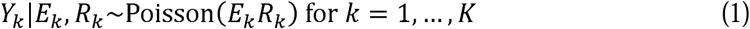

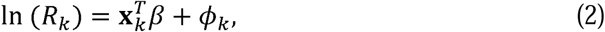

where the risk of mortality in area *k* is denoted by R_k_, x^T^is the matrix of covariates representing indicators of social disorganization, and the random effects are specified as <p = (<p_1_,…, <p_K_). The random effects allow for overdispersion and spatial correlation in the data after controlling for the social disorganization effects. Following Lee [71] and Lee and Mitchell [72] the S.CAR dissimilarity function in R was used to run the localized CAR model, including the *K x K* dissimilarity matrix. As applied to the present analysis, incorporating the matrix into the modeling scheme allowed us to identify the magnitude of the difference in EHI between two areas and incorporate the results into the model as a predictor of risk. In this model, the random effects in neighborhoods that share a border are modeled as correlated or conditionally independent, depending on whether EHI in the two bordering areas is similar (correlated random effects) or very different (conditionally independent) [73]. Spatial correlation is induced using a CAR prior for the random effects via the *K x K* neighborhood matrix **W**. We fit three separate boundary detection models: (1) a model without covariates; (2) a model with concentrated disadvantage, residential mobility, racial/ethnic diversity, and immigrant concentration; and (3) a final model that included the lagged variables for each covariate. Inference for all models was based on 10,000 post-burn-in and thinned Markov Chain Monte Carlo (MCMC) samples and model convergence was based on Geweke diagnostics. The residuals were assessed for spatial correlation using Moran’s I permutation test. Moran’s I measures the degree of spatial autocorrelation, indicating whether similar values are clustered or dispersed across a geographic area. A permutation test is used to assess if the observed Moran’s I value is statistically significant, which compares the observed value to a distribution of Moran’s I values generated under random spatial arrangements.

## Results

Between 2015 and 2023, the Cook County Medical Examiner investigated 2,226 deaths of children under 18. Table 1 presents the descriptive characteristics of the most (and least) deprived 10% of neighborhoods. By definition, the most deprived 10% of neighborhoods exhibited extreme levels of EHI compared to the least deprived 20% (most deprived: mean = 142, SD = 10; least deprived: mean = 74.4, SD = 2). Notably, the least deprived 10% of neighborhoods had less variability in EHI scores. On average, there were 1.59 child fatalities in the most deprived neighborhoods, resulting in a standardized mortality ratio (SMR) of 4.3 (Table 1). This indicates that child fatalities in the most deprived neighborhoods were 330% higher than expected if fatalities were randomly distributed throughout the study area. In contrast, the least deprived neighborhoods had an SMR of 0.16, indicating significantly fewer child fatalities than expected. These neighborhoods were also characterized by significantly higher levels of racial/ethnic homogeneity, concentrated disadvantage, and residential instability. Conversely, the most deprived neighborhoods had lower concentrations of immigrants and spent less on housing and transportation costs relative to income.

**Table 1.** Descriptive Characteristics of Key Study Variables.

| Characteristic | Least Deprived<br>10% of<br>Neighborhoods<br>(Q1, N = 132)<sup>1</sup> | Most Deprived<br>10% of<br>Neighborhoods<br>(Q10, N =<br>132)<sup>1</sup> | Diff.<sup>2</sup> | p-value<sup>2</sup> |
| --- | --- | --- | --- | --- |
| Economic Hardship & Inequality | 74.4 (2) | 142.0 (10) | -68 | <0.001 |
| Observed Child Deaths | 0.11 (0.34) | 1.59 (1.53) | -1.5 | <0.001 |
| Expected Child Deaths | 0.70 (0.49, 0.84) | 0.41 (0.26,<br>0.59) | 0.23 | <0.001 |
| Standardized Incidence Ratio<br>(SIR) | 0.16 (0.51) | 4.30 (6.20) | -4.1 | <0.001 |
| Residential Racial Segregation<br>(HHI) | 0.30 (0.14) | 0.14 (0.14) | 0.16 | <0.001 |
| Concentrated Disadvantage | -0.95 (0.30) | 1.69 (0.74) | -2.6 | <0.001 |
| Residential Instability | -1.35 (0.45) | 1.26 (0.52) | -2.6 | <0.001 |
| Immigrant Concentration | 0.04 (0.04) | 0.02 (0.03) | 0.02 | <0.001 |

<sup>1</sup>Mean (SD); Median (IQR)
<sup>2</sup>Welch Two Sample t-test
<sup>3</sup>CI = Confidence Interval

Moran’s I = 0.76 (*p* < .001) suggests a strong positive spatial autocorrelation, meaning that areas with similar EHI values are geographically proximate. Figure 2A illustrates the geographic distribution of EHI across census tracts in Cook County, visually showing how EHI values are clustered across the region. The figure also shows that many areas are characterized by differences in EHI, some of which appear stark. The histogram in Figure 2B shows a right-skewed distribution in EHI, meaning more neighborhoods are characterized by levels of EHI below the mean (100), fewer neighborhoods have levels above the mean, and a few have very extreme levels of EHI (> 140).

**Figure 2.**
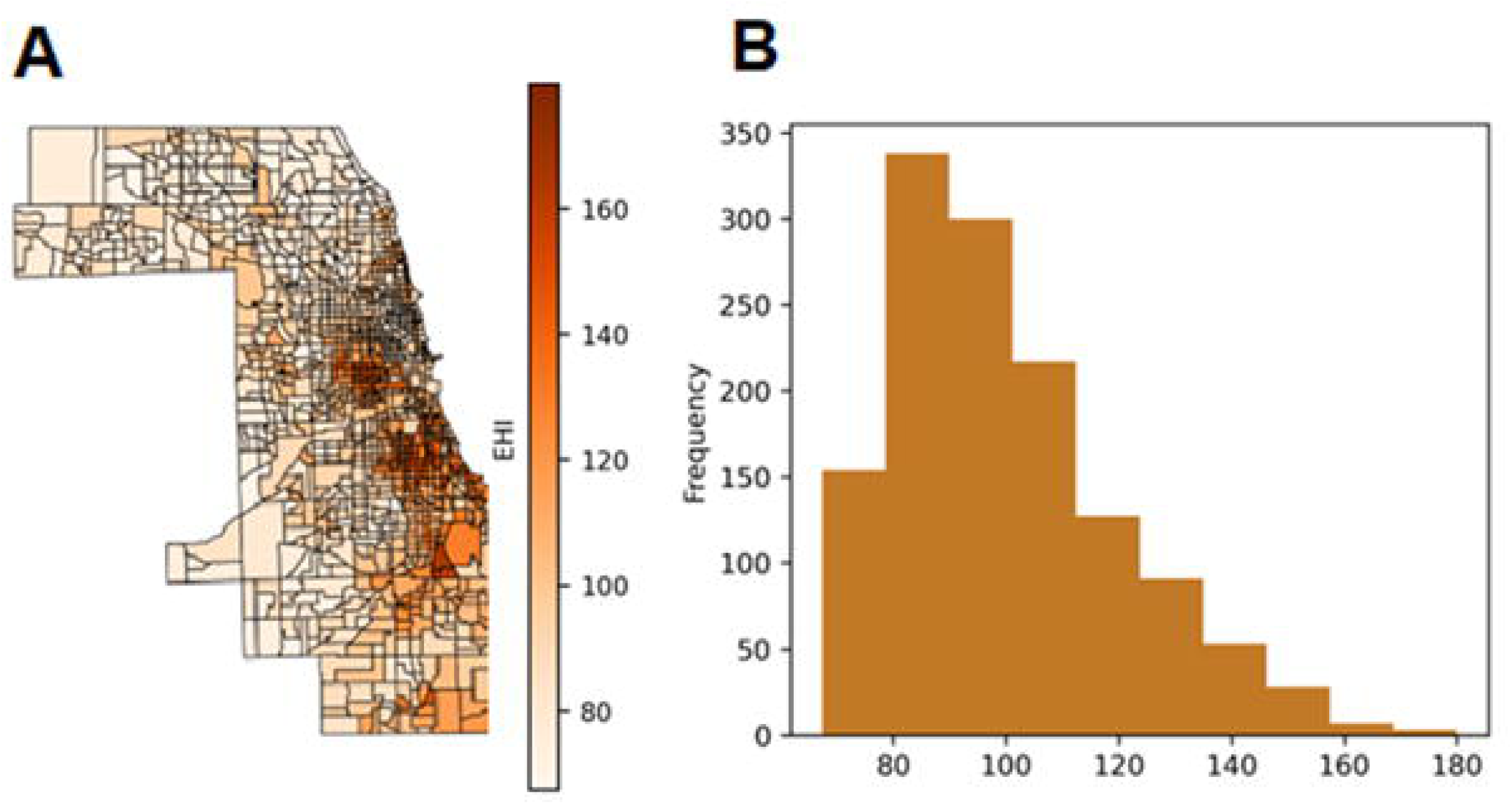
Distribution of Economic Hardship and Inequality. Geographic distribution of EHI (A; left); Histogram of EHI scores (B; right) for Cook County, Chicago.

Figure 3 shows the distribution of differences in the EHI across all pairs of census tracts. This analysis provides insight into whether the EHI distributions are similar or different between neighboring and non-neighboring tracts. The figure shows that the two distributions are dissimilar; neighboring tracts have *smaller wealth differences than* non-neighboring tracts, confirming the presence of spatial autocorrelation among neighbors. This means nearby tracts have more minor differences in EHI than non-neighboring tracts. A map randomization process allows us to detect significant differences at least as large as a random distribution. In Figure 4B, the result shows that the random distribution is more dispersed compared to the observed distribution of differences among nearby tracts. Despite evidence that EHI is highly clustered across the county, the spatial weights matrix detected 413 distinct boundaries where the difference in EHI deciles between adjacent tracts is at least four. We map these differences in Figure 3. The figure shows a localized spatial structure, with numerous distinct areas characterized by a high and low-risk border. This, in turn, suggests that boundaries are likely present in the risk surface, and their identification is the goal of this analysis.

**Figure 3.**
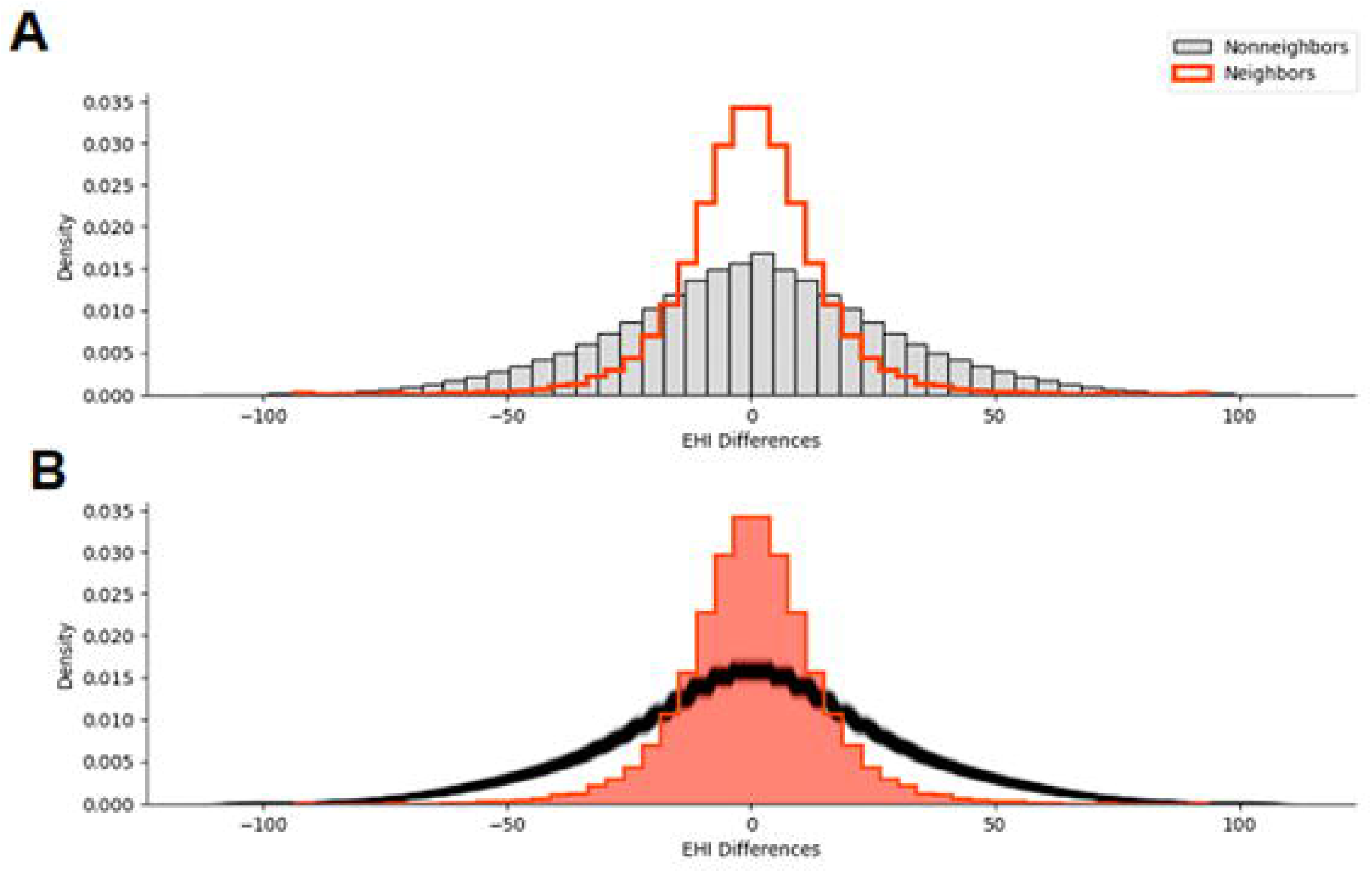
Neighborhood Differences in Economic Hardship and Inequality. Distribution of differences in the Economic Hardship and Inequality (EHI) across pairs of neighboring census tracts (A) and the randomization process used to detect significant differences (B). The peak of the histogram centered around zero represents no difference in EHI between tracts, meaning that two census tracts have the same level of EHI. The analysis shows that neighboring tracts are more likely to have few differences than non-neighboring tracts. In other words, neighboring tracts have similar levels of EHI compared to non-neighbors, and the result is significant.

**Figure 4.**
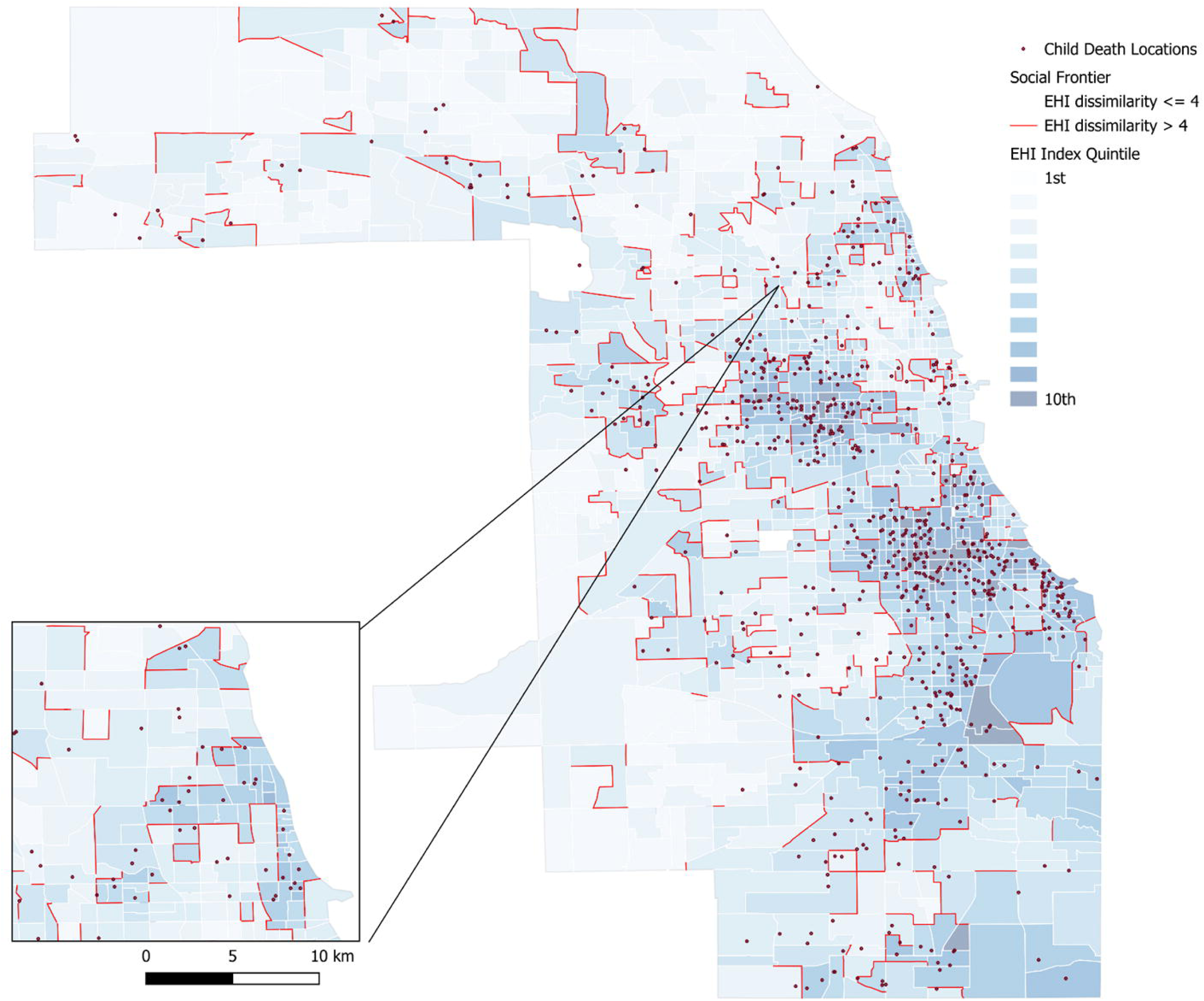
Boundary Detection Map. Map of Cook County Chicago overlaid with locations of child deaths and deciles of Economic Hardship and Inequality (EHI). Areas shaded darker blue represent higher levels of EHI. The red borders represent areas where the difference in EHI decile is > 4, indicating a spatial discontinuity.

The spatial dissimilarity model evaluates how economic inequality across neighborhood boundaries influences child mortality risk, progressing through four stages of model specification: an intercept-only model, a model incorporating social disorganization variables, and a fully lagged model. Table 2 summarizes the findings, including log risk surface estimates, ln (0_k_), the intercept, random effects terms, Geweke’s diagnostic for model convergence, and the threshold parameter (α). This threshold determines whether the dissimilarity metric effectively identifies boundaries in the random effects surface. In the intercept-only model (Model 1), focusing solely on EHI boundaries as the independent variable, the results show a 22.2% higher relative risk of child mortality at neighborhood boundaries with large EHI differences (exp^0.2002^ = 1.2216). The posterior mean and 95% credible interval of the regression parameter α lie completely above the threshold (α_min_), indicating that the EHI dissimilarity metric identifies statistically significant boundaries. In the second model (Model 2), four social disorganization variables—concentrated disadvantage, racial/ethnic diversity, immigrant concentration, and residential instability—are added. Controlling for these variables, concentrated disadvantage and residential instability remain significantly associated with higher child mortality risk, while neighborhoods with higher proportions of immigrant groups show a decreased risk. These findings suggest that social and economic neighborhood characteristics contribute to child mortality risk at boundaries. The third model (Model 3) extends the analysis by including spatially lagged versions of the four social disorganization variables to capture the broader spatial context’s influence on child mortality. Concentrated disadvantage remains significant, with a 29.2% increase in relative risk of child deaths for each standard deviation increase in residential instability, emphasizing its widespread spatial impact. This indicates that proximity to areas with high concentrated disadvantage significantly increases the risk of child fatalities, highlighting the effect of surrounding neighborhoods on child mortality.

**Table 2.** Results from the Spatial Discontinuity Models.

| **Table 2.** Results from the Spatial Discontinuity Models |  |  |  |  |  |  |
| --- | --- | --- | --- | --- | --- | --- |
|  | **Model 1** |  |  | **Model 2** |  | **Model 3** |
|  | Relative Risk | 95% Credible Interval | Relative Risk | 95% Credible Interval | Relative Risk | 95% Credible Interval |
| Intercept | -0.655 | -0.787, -0.535 | -0.398 | -0.695 - 0.140 | -0.429 | -3.059, 1.927 |
| $\tau^2$ | 1.664 | 1.231, 2.092 | 0.962 | 0.528, 1.447 | 0.584 | 0.327, 0.899 |
| HHI |  |  | -0.062 | -0.579, 0.582 | -0.102 | -1.029, 0.574 |
| Concentrated disadvantage |  |  | 0.244 | 0.113, 0.362 | -0.007 | -0.173, 0.141 |
| Residential instability |  |  | 0.291 | 0.149, 0.427 | 0.061 | -0.111, 0.226 |
| Immigrant concentration |  |  | -2.233 | -3.59, -0.946 | -1.872 | -4.240, 0.509 |
| HHI lag |  |  |  |  | 0.375 | -0.931, 1.721 |
| Concentrated disadvantage lag |  |  |  |  | 0.580 | 0.300, 0.833 |
| Residential instability lag |  |  |  |  | -0.186 | -0.472, 0.108 |
| Immigrant concentration lag |  |  |  |  | -1.919 | -5.249, 0.378 |
| **Boundary: EHI** | 0.20<br>0 | 0.174, 0.227 | 0.188 | 0.038, 0.229 | 0.193 | 0.091, 0.228 |

**Table notes.** EHI = Economic Hardship and Inequality; The boundary EHI quantifies the effect of living in an ‘social frontier’ on the relative risk of child fatality in a given census tract. HHI = Hirschman-Herfindahl Index is a measure of racial/ethnic segregation in a given census tract. The lag variables capture the influence of surrounding regions’ on that measure on the child death outcomes in the focal region.

Across all models, posterior medians, 95% credible intervals (crI), and minimum threshold values (α_min_=0.077), are reported for the economic inequality variable, confirming that the EHI dissimilarity metric consistently detects boundaries in the risk surface. Illustrating the intersection of economic inequality and child fatality risk, Figure 4 shows the overlay of the *K x K* symmetric matrix containing the posterior median for the set {w_kj_ |k∼J} onto the map of area-level economic inequality where k∼J denotes areas that share a common border.

In Figure 5A, the 413 boundaries are represented as solid green lines, with line thickness corresponding to higher probabilities of child mortality risk. The most significant boundaries are located at the divisions between regions in the Northeast and Midwest sections of the county, with the most prominent boundary occurring between the central-western and northeastern parts of Chicago. The boundaries identified in the analysis represent 10.6% of all borders between census tracts in the study region, indicating that a relatively small but meaningful portion of the census tract boundaries are associated with sharp contrasts in economic inequality (Figure 5B). Even after controlling for multiple social disorganization factors and their spatially lagged effects, the influence of economic inequality boundaries on child mortality remains significant. These findings underscore the role of stark EHI contrasts at neighborhood boundaries in increasing child mortality risk, independent of internal neighborhood characteristics and broader spatial processes (Figure 5C) [51].

**Figure 5.**
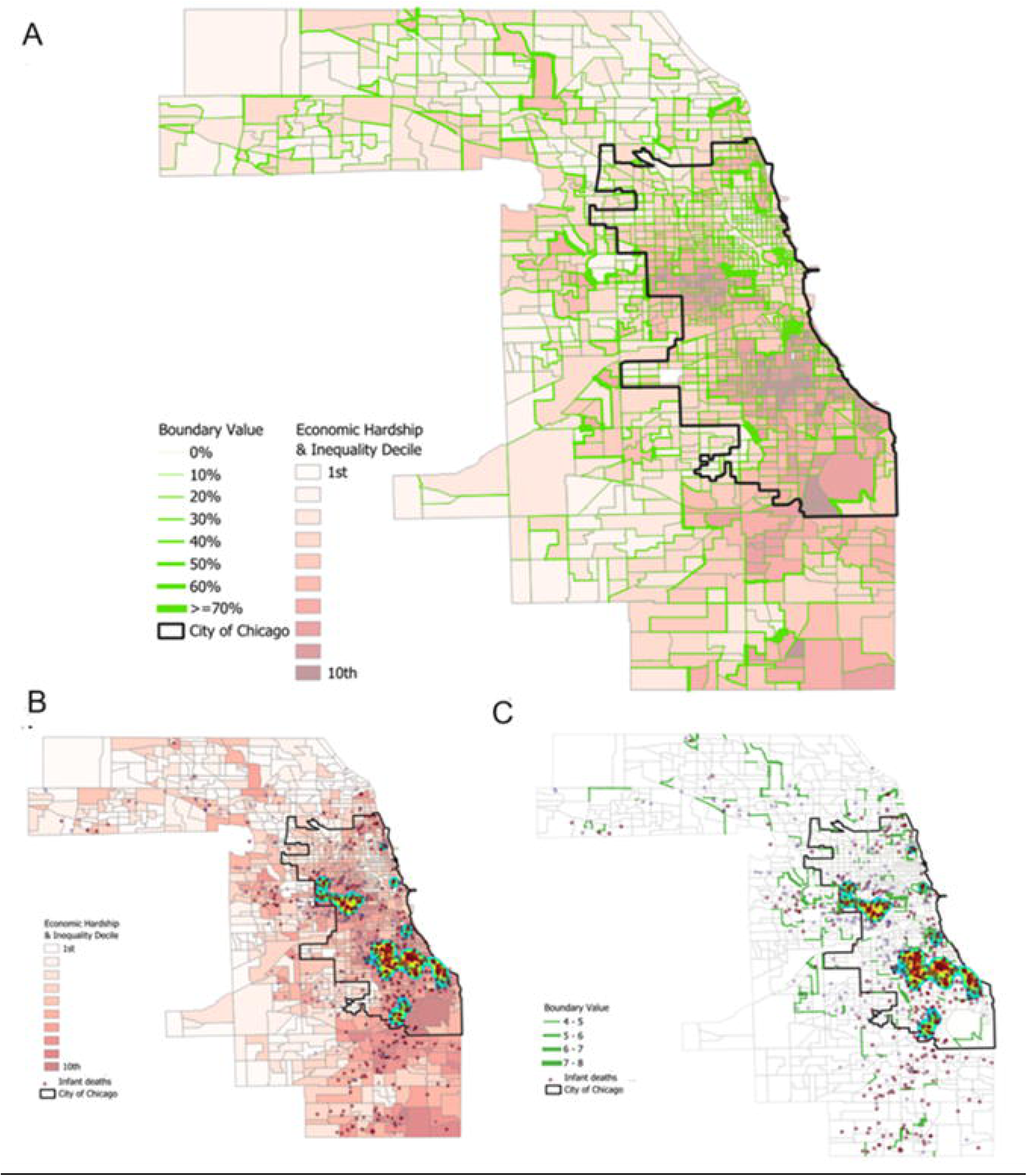
Spatial Frontiers and Child Mortality. (A) Economic Hardship and Inequality (EHI; shaded red) by decile and boundary value of EHI (shaded green). The thickness of the boundary reflects higher inequality across the EHI boundary; (B) EHI by decile (shaded red) and location (represented by the dots) and density of infant deaths; (C) Boundary locations of social frontier representing areas where differences in EHI > 4 across census tracts (green lines) and locations of child deaths (represented by the dots).

## Discussion

Rather than focus on single spatial units as political boundaries, this study sought to identify spatial discontinuities in EHI defined by extreme differences in socioeconomic inequality and its impact on child mortality risk. Until now, no study has previously captured the localized spatial structure of EHI and its influence on child mortality risk in bordering neighborhoods [56]. The analysis used in this paper identified 413 neighborhood boundaries (10% of the total boundaries in Cook County) characterized by sharp changes in EHI [52]. These areas which we refer to as social frontiers are associated with a 22% higher relative risk of child mortality. Further, the increased risk persisted even after accounting for key indicators of social disorganization, including racial segregation, concentrated disadvantage, residential mobility, and immigrant concentration. The results of this study align with previous research demonstrating how social and spatial factors shape local opportunities based on extreme differences in socioeconomic status, which in turn affect the health outcomes of children living in urban areas.^53^

Descriptive results showed that the most deprived neighborhoods have substantially more child deaths than would be expected if the distribution were randomly distributed in the county, indicating a higher relative risk in some neighborhoods. This risk, quantified by the SIR, is 330% greater in the most deprived 10% of neighborhoods and is consistent with previous research demonstrating socioeconomic gradients in child health [54–55]. The spatial distribution of EHI in Cook County neighborhoods, defined by census tracts, revealed stark differences, providing further evidence of spatial heterogeneity, meaning that EHI is not evenly distributed across the region. In other words, areas of extreme advantage are much more concentrated in certain areas of the city. Further, neighborhoods that are closer together are characterized by smaller differences in EHI compared to neighborhoods that are farther away. Despite EHI’s strong spatial clustering, we found substantially less variability in the least deprived 10% of neighborhoods compared to the most deprived 10%. In addition, we identified a localized spatial structure where extreme differences in EHI are characterized by latent discontinuities in the risk surface. These findings suggest that the geographic concentration of advantage contributes more to inequality in child mortality than the concentration of EHI, given the increased risk of child mortality in social frontiers.

Our findings highlight the interconnectedness and interdependency across socioeconomic divides in different regions. Even after controlling for multiple measures of social disorganization, the impact of economic inequality boundaries remained significant. Specifically, we found that in social frontiers, the relative risk of child mortality is 22% higher net racial/ethnic diversity, concentrated disadvantage, residential mobility, and immigrant concentration. One explanation is provided by Kramer, who argues that highly segregated boundaries “orient and abet other inequalities by reifying differences between otherwise similar urban spaces ”[47]. Similarly, the number of social frontiers uncovered in this study provide evidence of relative resource deprivation across Cook County [57]. The study found that areas surrounded by neighborhoods of higher concentrated disadvantage and residential instability have increased risks of child mortality, further indicating a spatial spillover effect. In these areas, relative resource deprivation may affect a family’s ability to meet its basic needs with the resources available in the community [58]. As a result, neighborhoods with elevated levels of disadvantage and instability not only have higher mortality rates within their boundaries but also contribute to higher mortality rates in neighboring areas. These stark contrasts in EHI reflect economic disparities and have direct consequences for child health, illustrating how location can dictate access to resources and structure opportunities across socioeconomic divides.

### Implications for policy and practice

The results of this study hold several public policy implications relevant to child health outcomes. Considering our results, policies aimed at reducing economic inequality rather than poverty or increasing income can play a vital role in mitigating child mortality risks. For example, cash transfer programs that reduce poverty, improve economic autonomy, child nutrition, and health service utilization, are associated with significant reductions in mortality among children under five years of age and women [74]. However, while such programs reduce poverty, they have been shown to have a more limited impact in reducing inequality [75]. On the other hand, mixed-income housing developments may effectively address inequality with other interventions that reduce prejudice and intergroup competition, facilitate civic participation, and support social well-being in built environments [76]. There are other effective way to address economic inequality by focusing on neighborhood transformation through place-based initiatives [77], such as the Dudley Street Initiatives Promise Neighborhoods Program in Boston. Place-based strategies deliver comprehensive social services—healthcare, education, child welfare— and can help families living in more deprived areas receive the same level of support as those in more affluent areas. By addressing neighborhood inequalities directly, these policies can provide children with spatial access to quality education, healthcare providers, and safer environments, thereby indirectly minimizing child mortality risk. By focusing on EHI, these interventions will both reduce child mortality and foster long-term resilience in the most disadvantaged neighborhoods.

Social frontiers should be public health priority areas where interventions are focused on bridging the gap in EHI between affluent and deprived areas through infrastructure and community development initiatives. These initiatives include affordable community centers and healthcare facilities that offer multifaceted services conditioned by local needs. To address spatial discontinuities in EHI, interventions to reduce child mortality risk should focus on localized opportunity structures rather than administratively defined areas [78]. This includes services targeted to areas of relative resource deprivation, i.e., “social frontiers” that serve both sides of the divide but are specifically designed to reduce the stark inequality at the boundary. Implementing mobile health clinics or social services that specifically target neighborhoods at socioeconomic boundaries can improve access to healthcare in disadvantaged areas. By targeting areas of concentrated disadvantage, we can directly confront the socioeconomic divides that contribute to higher child mortality rates.

### Limitations

This study underscores the critical role of neighborhood environments in shaping health and socio-economic inequalities. Our conceptual and methodological approach highlights the importance of spatial factors in shaping policies to improve child well-being by calling attention to neighborhood boundaries and economic disparities as critical elements in reducing child mortality, particularly in urban areas. Nevertheless, our study is not without limitations. First, the use of area-level data, such as census tracts, may obscure finer-scale variations within neighborhoods, limiting the ability to capture small-scale dynamics that influence child mortality. Additionally, the model assumes linear relationships between socioeconomic inequality (EHI) and child mortality, which may not fully capture more complex interactions. There is also the potential for unmeasured confounding factors that we did not consider, such as healthcare access or local policies, which could influence socioeconomic conditions and child mortality rates. The cross-sectional nature of the data limits the ability to account for temporal dynamics, and the findings may not generalize beyond Cook County, Illinois. Although our results were robust to alternative specifications for the EHI dissimilarity metric, using other indicators of socioeconomic deprivation may result in different results. These limitations should be considered when interpreting the findings, and future research would benefit from replicating our results to provide more insight into the impact of broadly defined inequality on child mortality.

## Conclusion

In summary, spatial spillover in child mortality across neighborhoods is a complex phenomenon influenced by many factors. Understanding this can help develop targeted interventions to address these disparities and improve child health outcomes [1]. The study underscores the importance of targeted interventions considering different neighborhoods’ specific characteristics that reduce economic inequality, including secure housing and accessibility to transportation and jobs, paying particular attention to how it plays out over geographic space [3].

## Data Availability

All data are publicly available from the Cook County Medical Examiner Case Archive (https://datacatalog.cookcountyil.gov/Public-Safety/Medical-Examiner-Case-Archive/cjeq-bs86/data), the U.S. Census Bureau (https://www.census.gov/data.html), and the Area Deprivation Index from the Neighborhood Atlas (https://www.neighborhoodatlas.medicine.wisc.edu/)

